# Long-Term Coronary Microvascular and Cardiac Dysfunction Following Severe COVID-19 Hospitalization

**DOI:** 10.1101/2024.11.14.24317343

**Authors:** Rebecka Steffen Johansson, Daniel Loewenstein, Klara Lodin, Judith Bruchfeld, Michael Runold, Marcus Ståhlberg, Hui Xue, Peter Kellman, Kenneth Caidahl, Henrik Engblom, Jannike Nickander

## Abstract

**Background:** Coronavirus disease 2019 (COVID-19) can lead to long-term cardiopulmonary symptoms and is associated to coronary microvascular dysfunction (CMD). However, long-term data on CMD following severe COVID-19 are lacking.

**Objective:** To determine long-term left ventricular (LV) function and presence of CMD after severe COVID-19, utilizing cardiovascular magnetic resonance (CMR) and stress perfusion mapping.

**Methods:** Hospitalized COVID-19 patients underwent CMR at 10 months follow-up (1.5T Aera, Siemens Healthineers) including cine imaging, native T1 and T2, extracellular volume, and adenosine stress perfusion mapping. Clinical data were obtained from patient records. Patients were compared to volunteers without symptomatic ischemic heart disease (IHD).

**Results:** COVID-19 patients (n=37, age 56±12 years, 76% male) and volunteers (n=22, age 51±13 years, 55% male, p=ns for both) were included. COVID-19 patients demonstrated reduced stress perfusion (2.8±0.81 vs 3.4±0.69 ml/min/g, *p*=0.003), impaired global longitudinal strain (GLS, −17±2 vs −19±2 %, *p*=0.003) and global circumferential strain (GCS, −16±3 vs −19±3 %, *p*=0.001). There were no differences in stress perfusion or myocardial perfusion reserve between COVID-19 patients with or without cardiovascular risk factors or cardiac symptoms.

**Conclusion:** COVID-19 patients exhibit long-term reduced stress perfusion indicating CMD, and impaired LV function by GLS and GCS. Lack of variation in stress perfusion between patients with and without cardiovascular risk factors suggests that CMD may be a consequence of severe COVID-19, warranting further investigation to elucidate mechanisms, and guide potential therapeutic interventions.

## Introduction

Coronavirus disease 2019 (COVID-19) primarily presents with respiratory symptoms of varying severity, although extrapulmonary manifestations such as cardiac complications are common, and importantly, correlate with disease severity and risk of mortality^1^. Long-term follow-up studies reveal heightened risk of arrhythmias, ischemic and non-ischemic heart disease, heart failure, perimyocarditis and thromboembolic events among both non-hospitalized and hospitalized COVID-19 patients, with highest risk in those requiring intensive care during the first waves of the pandemic^2^. Moreover, in long-Covid^3^, also termed post-acute COVID-19 syndrome (PACS)^4^, it is common with cardiopulmonary symptoms such as dyspnea, palpitations, chest pain and fatigue, which impair quality of life and functional capacity^5–7^. The underlying pathophysiological mechanisms are not yet fully understood but may stem from myocardial injury sustained during acute COVID-19 due to hypoxia, systemic hyper-inflammation, hypercoagulability and direct viral invasion of endothelial cells and cardiomyocytes^8,9^. Notably, microvascular dysfunction across various vascular beds, including the coronary circulation, has been documented following COVID-19^10–13^. Coronary microvascular dysfunction (CMD) may manifest as chest pain with or without obstructive coronary artery disease (CAD), and traditional cardiovascular risk factors are associated with both CMD and risk of severe COVID-19^14^. CMD can be detected using quantitative cardiovascular magnetic resonance (CMR) adenosine stress perfusion mapping^15^. Moreover, comprehensive multiparametric CMR may characterize cardiac anatomy, function, and tissue properties^16^. Despite the critical need for understanding long-term cardiovascular implications in PACS, long-term data on critically ill patients are limited. Thus, the aim of this study was to evaluate presence of CMD in patients hospitalized due to severe COVID-19 in long-term follow-up.

## Methods

### Study population

Patients were identified from the prospective study “Follow-up of patients with severe COVID-19” (UppCov), aiming to comprehensively assess long-term outcome following hospitalization due to severe COVID-19, at Karolinska University Hospital, Stockholm, Sweden^17^. Severe COVID-19 was defined as respiratory failure requiring ventilatory support and/or oxygen therapy (oxygen flow ≥5 L/minute). In total, 40 COVID-19 patients with and without cardiac involvement during hospitalization were included in the present study from November 2020 to February 2021 for CMR at approximately 10 months follow-up. Cardiac involvement was defined as high-sensitive troponin T (hs-TnT)>14 ng/L and/or pulmonary artery pressure (PAP)>34 mmHg. Exclusion criteria were general contraindications for adenosine stress CMR, including claustrophobia, pacemaker or CMR-incompatible metal implants, severe asthma or severe chronic obstructive pulmonary disease (COPD), high degree atrioventricular block and renal failure (estimated glomerular filtration rate, eGFR<30 mL/minute/1.73 m^2^). Patients with known angina pectoris, previous myocardial infarction (MI), previous coronary artery bypass grafting and/or percutaneous coronary intervention, stroke, heart failure, aortic stenosis or arrhythmias including atrial fibrillation were excluded. Historical volunteers with similar age and sex, without symptomatic ischemic heart disease (IHD), were included for comparison^18,19^. COVID-19 patients were invited to clinical follow-up 244 [214-288] days post-discharge. Residual respiratory symptoms were evaluated using the COPD assessment test (CAT) ^20^and the modified medical research council dyspnea scale (mMRC)^21^. Quality of life was assessed using the EQ visual analogue scale (EQ-VAS)^22^. Clinical data including previous diseases and cardiovascular risk factors, medications, details regarding the hospital stay and clinical follow-up, were obtained from medical records and by interviews. All procedures were granted ethical approval by the Swedish Ethical Review Authority (Dnr 2021-03293, 2022-0695, 2020-02397). All participants provided written informed consent.

### Image acquisition

Patients and volunteers underwent the same CMR protocol. CMR was performed supine with a 1.5 T Aera® scanner (Siemens Healthineers, Erlangen, Germany), a phased-array 18-channel body matrix coil and a spine matrix coil. Hematocrit and blood creatinine were determined prior to CMR. Full coverage retrospective electrocardiographic (ECG)-gated balanced steady state free precession (bSSFP) cine imaging was acquired in short-axis and three-long axis slices. Typical parameters were flip angle (FA) 68°, pixel size 1.4×1.9 mm^2^, slice thickness 8.0 mm, echo time (TE)/repetition time (TR) 1.19/37.05 ms, matrix size 256×144 and field of view (FOV) 360×270 mm^2^.

Quantitative perfusion maps were acquired in three short-axis slices using first pass perfusion imaging, during adenosine infusion (140 µg/kg/min or increased according to clinical routine to 210 µg/kg/min (Adenosin, Life Medical AB, Stockholm, Sweden)) and in rest, during injection of intravenous contrast (0.05 mmol/kg, gadobutrol, Gadovist, Bayer AB, Solna, Sweden). Subjects were asked to abstain from caffeine for 24 hours and nicotine for 12 hours prior to CMR. Adenosine response was assessed based om symptoms and heart rate response. Contrast and adenosine were administered in separate cannulas. Perfusion maps were computed using the distributed tissue exchange model ^23^ and generated using the Gadgetron inline perfusion mapping software^24^. Typical parameters were bSSFP single shot readout, FA 50°, slice thickness 8.0 mm, TE/TR 1.04/2.5 ms, bandwidth 1085 Hz/pixel, FOV 360×270 mm^2^, saturation delay/trigger delay 95/40 ms.

Native T2 maps were acquired in three or five short-axis slices using a T2-prepared sequence (Siemens MyoMaps product sequence). Typical parameters were FA 70°, pixel size 1.4×1.4 mm^2^, slice thickness 8.0 mm, acquisition window 700 ms, TE/TR 1.06/2.49 ms, matrix size 144×256 mm^2^. Native T1 maps were acquired in three or five short-axis slices using an ECG-gated modified look-locker inversion recovery (MOLLI) ^25^5s(3s)3s research sequence. Typical parameters were end-diastole single shot SSFP, FA 35°, pixel size 1.4×1.4 mm^2^, slice thickness 8.0 mm, imaging duration 167 ms, TE/TR 1.12/2.7 ms, matrix size 256×144 and FOV 360×270 mm^2^. Post-contrast T1 maps were acquired following a contrast bolus (0.2 mmol/kg, gadobutrol) with the same image positions as the native T1 maps. Extracellular volume (ECV) maps were generated from native and post-contrast T1 maps and calibrated by hematocrit^26^. Furthermore, post-contrast late gadolinium enhancement (LGE) images were acquired in short-axis and three long-axis slices using a free breathing phase-sensitive inversion recovery (PSIR) sequence with bSSFP single shot readout. Typical parameters were image matrix 256×156, voxel size 1.3×1.3×7 mm^3^, slice thickness 8 mm, FOV 340×276 mm, TR 8.25 ms, TE 3.4 ms and FA 50°.

### Image analysis

Images were anonymized and analyzed offline using Segment® (vers 2.7, Medviso AB, Lund, Sweden)^27,28^. Left ventricular (LV) volumes and mass were acquired using automatic segmentation of the cine short-axis stack in end-diastole and end-systole, with manual corrections. LV volumes and mass were indexed to body surface area (BSA), calculated with the Mosteller formula^29^. LV global longitudinal strain (GLS) and global circumferential strain (GCS) were acquired using the feature-tracking module in Segment, following delineation in end-diastole of the LV endo- and epicardial borders in the cine long-axis slices and short-axis stack. Native T1, native T2, ECV and perfusion maps were analyzed by manually delineating epi- and endocardial contours in respective short-axis stack. To avoid contamination from blood pool and adjacent tissues, a 10% erosion margin was set for endo- and epicardial borders. Segmental values were acquired in a 16-segment LV model^30^. Inter-observer analysis was performed in all patients by two separate observers, and intra-observer analysis was performed in 10 patients by one observer, by repeated analysis of the short-axis cine stack, native T1, native T2, ECV, and perfusion maps.

### Statistical analysis

Normality was assessed with the Shapiro-Wilk test. Continuous data were presented as mean ± standard deviation (SD) or as median [interquartile range], categorical data as frequencies (percentages). Global native T1, native T2, ECV and perfusion values were acquired by averaging segmental values. Myocardial perfusion reserve (MPR) was calculated as stress perfusion divided by rest perfusion. Rest perfusion was adjusted for rate pressure product (RPP), resting heart rate multiplied by resting systolic blood pressure. Patients and volunteers were compared with the independent t-test, Mann Whitney U-test or Fisher’s exact test, as appropriate. Intra- and interobserver agreement for LV ejection fraction (LVEF), GLS, GCS, global native T1, native T2, ECV, rest and stress perfusion and MPR, were calculated as intraclass correlation coefficient (ICC), using two-way random effects with absolute agreement. ICC ranged 0.72-1.00 (*p*<0.05 for all), Table 1 in Appendix. Microsoft Excel (version 16.6, Microsoft, Redmond, Washington, USA) and IBM SPSS Statistics (version 28, SPSS Statistics, IBM, New York, USA) were used for statistical analysis. The significance level was defined as *p*<0.05 in all statistical tests.

**Table 1.**
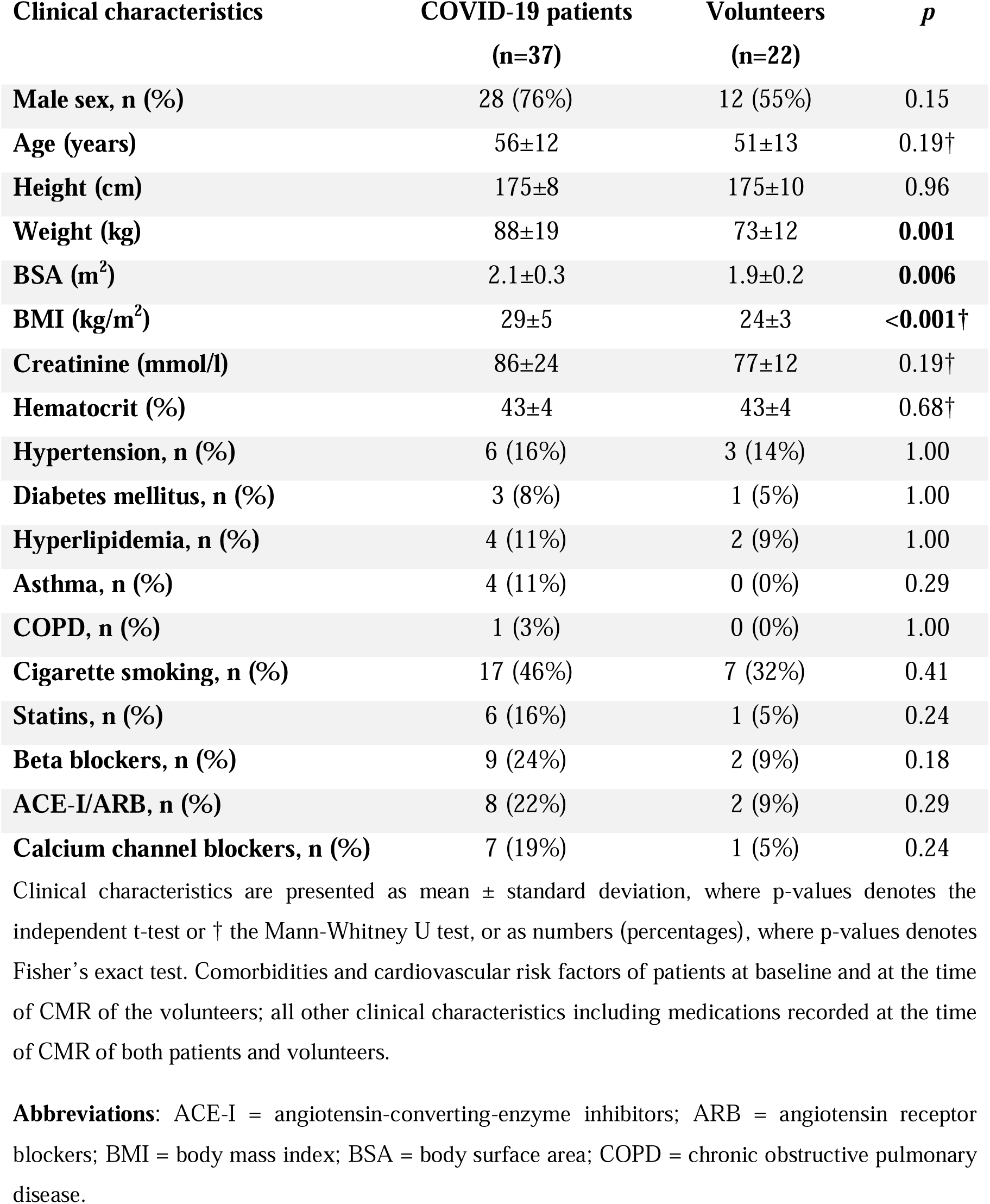
Clinical characteristics of the COVID-19 patients and volunteers.

| Clinical characteristics | COVID-19 patients<br>(n=37) | Volunteers<br>(n=22) | *p* |
| --- | --- | --- | --- |
| Male sex, n (%) | 28 (76%) | 12 (55%) | 0.15 |
| Age (years) | 56±12 | 51±13 | 0.19† |
| Height (cm) | 175±8 | 175±10 | 0.96 |
| Weight (kg) | 88±19 | 73±12 | **0.001** |
| BSA (m<sup>2</sup>) | 2.1±0.3 | 1.9±0.2 | **0.006** |
| BMI (kg/m<sup>2</sup>) | 29±5 | 24±3 | **<0.001†** |
| Creatinine (mmol/l) | 86±24 | 77±12 | 0.19† |
| Hematocrit (%) | 43±4 | 43±4 | 0.68† |
| Hypertension, n (%) | 6 (16%) | 3 (14%) | 1.00 |
| Diabetes mellitus, n (%) | 3 (8%) | 1 (5%) | 1.00 |
| Hyperlipidemia, n (%) | 4 (11%) | 2 (9%) | 1.00 |
| Asthma, n (%) | 4 (11%) | 0 (0%) | 0.29 |
| COPD, n (%) | 1 (3%) | 0 (0%) | 1.00 |
| Cigarette smoking, n (%) | 17 (46%) | 7 (32%) | 0.41 |
| Statins, n (%) | 6 (16%) | 1 (5%) | 0.24 |
| Beta blockers, n (%) | 9 (24%) | 2 (9%) | 0.18 |
| ACE-I/ARB, n (%) | 8 (22%) | 2 (9%) | 0.29 |
| Calcium channel blockers, n (%) | 7 (19%) | 1 (5%) | 0.24 |

Clinical characteristics are presented as mean ± standard deviation, where p-values denotes the independent t-test or † the Mann-Whitney U test, or as numbers (percentages), where p-values denotes Fisher's exact test. Comorbidities and cardiovascular risk factors of patients at baseline and at the time of CMR of the volunteers; all other clinical characteristics including medications recorded at the time of CMR of both patients and volunteers.
**Abbreviations:** ACE-I = angiotensin-converting-enzyme inhibitors; ARB = angiotensin receptor blockers; BMI = body mass index; BSA = body surface area; COPD = chronic obstructive pulmonary disease.

## Results

### Hospitalization due to COVID-19

COVID-19 patients were hospitalized 37 [20-54] days, from March to September of 2020. At presentation, 35 (95%) had fever, 34 (92%) had cough, 32 (86%) had dyspnea, 14 (38%) had myalgia, 13 (35%) had gastrointestinal symptoms and 2 (5%) had anosmia. Acute respiratory distress syndrome (ARDS) was diagnosed in 30 (81%) and graded as mild 2 (7%), moderate 6 (20 %) and severe 22 (73 %). Mechanical ventilation was required in 28 (76%) and 5 (14%) were placed on extracorporeal membrane oxygenation (ECMO). Continuous renal replacement therapy (CRRT) was utilized in 5 (14%), however all had eGFR>30 mL/minute/1.73 m^2^ at follow-up CMR. During hospitalization, 33 (89%) had hs-TnT>14 ng/L and 11 (30%) had PAP>34 mmHg – all but one of patients with elevated PAP also had elevated hs-TnT. Peak median hs-TnT during hospitalization was 51 [23-174] ng/L and peak median PAP was 50 [40-53] mmHg. Pulmonary embolism (PE) was present in 9/11 patients with elevated PAP, and in total 13 (35%) suffered PE during hospitalization. Additionally, 13 (35%) patients had arrhythmias, 11 (30%) had bacterial pneumonia and 13 (35%) had sepsis during hospitalization.

### Clinical characteristics and follow-up

Clinical characteristics are presented in Table 1. Although age and sex did not differ, COVID-19 patients had greater weight (88±19 vs 73±12 kg, *p*=0.001) and body mass index (BMI; 29±5 vs 24±3 kg/m^2^, *p*<0.001) compared to volunteers. No one in the study had a history of angina pectoris, MI, aortic stenosis, heart failure or arrhythmias and no one had severe asthma or COPD at follow-up CMR. There were no other differences between patients and volunteers in cardiovascular risk factors and medications. Clinical follow-up was performed 244 [214-288] days post-discharge. At follow-up, persistent cough was reported in 10 (27%), dyspnea in 16 (43%), chest pressure in 7 (19%) and fatigue in 13 (35%). Mean CAT score was 10±7 and median mMRC score was 1 [1-1.75] (data missing for 6 patients). Mean EQ VAS score was 72±18 (data missing for 7 patients). At clinical follow-up, 11 (44%) of the 25 patients that worked prior to hospitalization were still on sick leave; 12 patients were retirees.

### CMR at follow-up

CMR was performed 292 [203-367] days following discharge, or at approximately 10 months. Due to poor image quality or contraindications to stress CMR, three COVID-19 patients were excluded, rendering 37 patients included. CMR findings are presented in Table 2. Native T2 maps were not obtained in two COVID-19 patients due to operator dependency. COVID-19 patients had worse GLS (−17±2 vs −19±2%, *p*=0.003) and GCS (−16±3 vs −19±3%, *p*=0.001) compared to volunteers. There were no differences in LV mass or volumes, global native T1, native T2, or ECV. Minimal LGE was found in four COVID-19 patients; two had LGE consistent with a prior MI and two had LGE consistent with prior myocarditis. Due to limited scarring (<1 segment), the patients were included in the analysis.

**Table 2.**
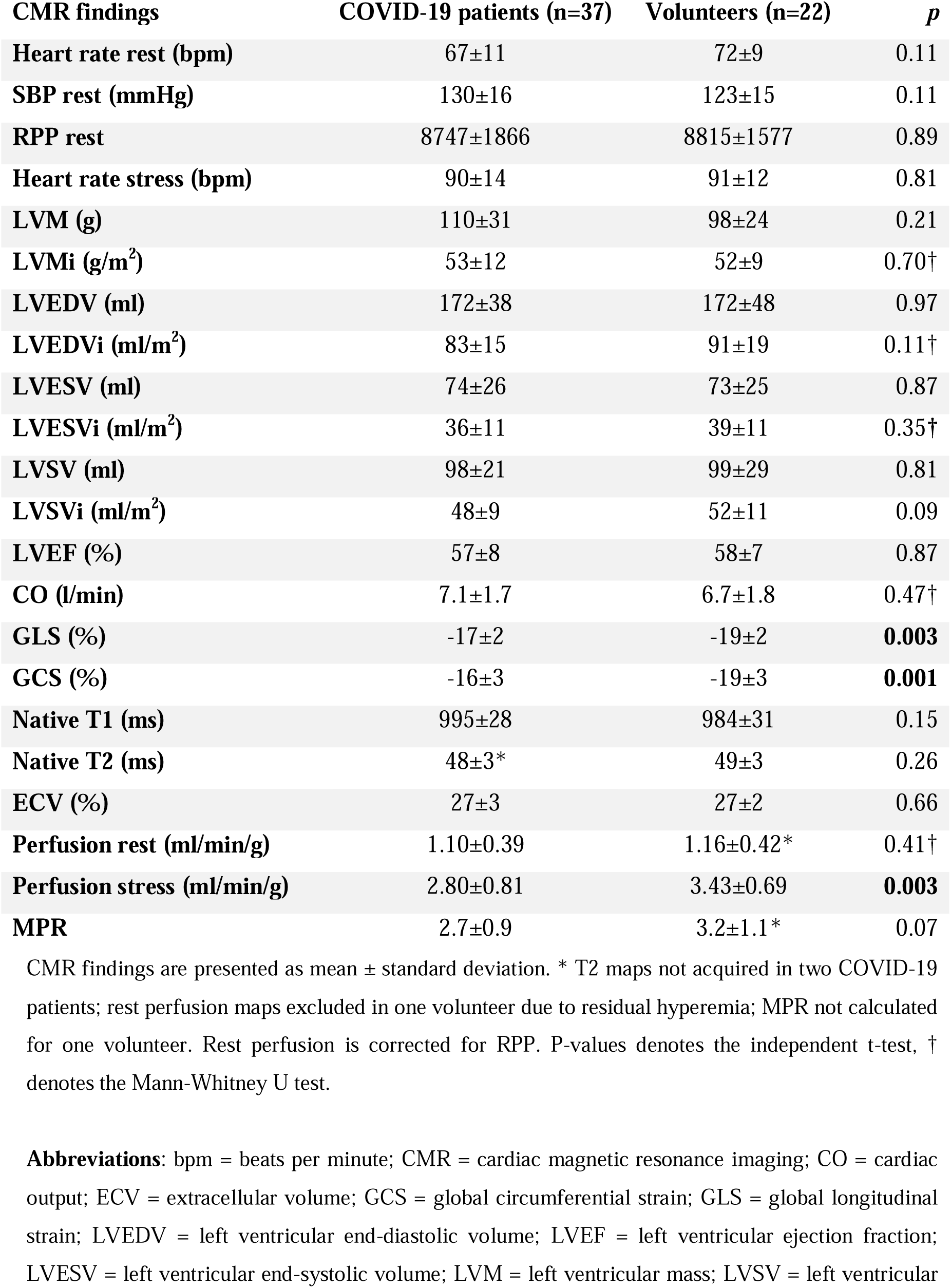
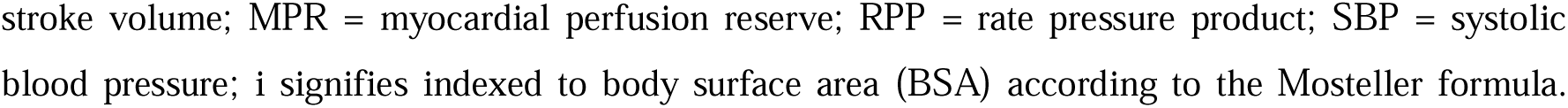
CMR findings of the COVID-19 patients and volunteers.

| CMR findings | COVID-19 patients (n=37) | Volunteers (n=22) | *p* |
| --- | --- | --- | --- |
| Heart rate rest (bpm) | 67±11 | 72±9 | 0.11 |
| SBP rest (mmHg) | 130±16 | 123±15 | 0.11 |
| RPP rest | 8747±1866 | 8815±1577 | 0.89 |
| Heart rate stress (bpm) | 90±14 | 91±12 | 0.81 |
| LVM (g) | 110±31 | 98±24 | 0.21 |
| LVMi (g/m<sup>2</sup>) | 53±12 | 52±9 | 0.70† |
| LVEDV (ml) | 172±38 | 172±48 | 0.97 |
| LVEDVi (ml/m<sup>2</sup>) | 83±15 | 91±19 | 0.11† |
| LVESV (ml) | 74±26 | 73±25 | 0.87 |
| LVESVi (ml/m<sup>2</sup>) | 36±11 | 39±11 | 0.35† |
| LVSV (ml) | 98±21 | 99±29 | 0.81 |
| LVSVi (ml/m<sup>2</sup>) | 48±9 | 52±11 | 0.09 |
| LVEF (%) | 57±8 | 58±7 | 0.87 |
| CO (l/min) | 7.1±1.7 | 6.7±1.8 | 0.47† |
| GLS (%) | -17±2 | -19±2 | **0.003** |
| GCS (%) | -16±3 | -19±3 | **0.001** |
| Native T1 (ms) | 995±28 | 984±31 | 0.15 |
| Native T2 (ms) | 48±3* | 49±3 | 0.26 |
| ECV (%) | 27±3 | 27±2 | 0.66 |
| Perfusion rest (ml/min/g) | 1.10±0.39 | 1.16±0.42* | 0.41† |
| Perfusion stress (ml/min/g) | 2.80±0.81 | 3.43±0.69 | **0.003** |
| MPR | 2.7±0.9 | 3.2±1.1* | 0.07 |

CMR findings are presented as mean ± standard deviation. \* T2 maps not acquired in two COVID-19 patients; rest perfusion maps excluded in one volunteer due to residual hyperemia; MPR not calculated for one volunteer. Rest perfusion is corrected for RPP. P-values denotes the independent t-test, † denotes the Mann-Whitney U test.
**Abbreviations:** bpm = beats per minute; CMR = cardiac magnetic resonance imaging; CO = cardiac output; ECV = extracellular volume; GCS = global circumferential strain; GLS = global longitudinal strain; LVEDV = left ventricular end-diastolic volume; LVEF = left ventricular ejection fraction; LVESV = left ventricular end-systolic volume; LVM = left ventricular mass; LVSV = left ventricular
stroke volume; MPR = myocardial perfusion reserve; RPP = rate pressure product; SBP = systolic blood pressure; i signifies indexed to body surface area (BSA) according to the Mosteller formula.

#### Quantitative myocardial perfusion

Stress perfusion was lower in COVID-19 patients compared to volunteers (2.8±0.81 vs 3.4±0.69 ml/min/g, *p*=0.003) but there was no difference in rest perfusion, Figure 1. There was a trend towards lower MPR in COVID-19 patients (2.7±0.9 vs 3.2±1.1, *p*=0.07). Rest perfusion maps were excluded in one volunteer due to residual hyperemia. Representative examples of perfusion maps of a COVID-19 patient with suspected CMD and a volunteer are presented in Figure 3. Stress and rest perfusion, and MPR, did not differ between female and male COVID-19 patients, nor between COVID-19 patients with or without a history of hypertension, hyperlipidemia, diabetes mellitus or cigarette smoking (*p*>0.05 for all, data not shown). COVID-19 patients reporting chest pressure at follow-up had higher MPR compared to patients without (3.3±1.2 vs 2.5±0.7, *p*=0.03), driven by non-significantly higher stress perfusion and lower rest perfusion (3.11±0.68 vs 2.72±0.82, *p*=0.26, and 0.99±0.25 vs 1.12±0.41 ml/min/g, *p*=0.78). Otherwise, there were no differences in perfusion or MPR between COVID-19 patients with or without chest pressure, dyspnea, or fatigue at follow-up (*p*>0.05 for all, data not shown).

**Figure 1.**
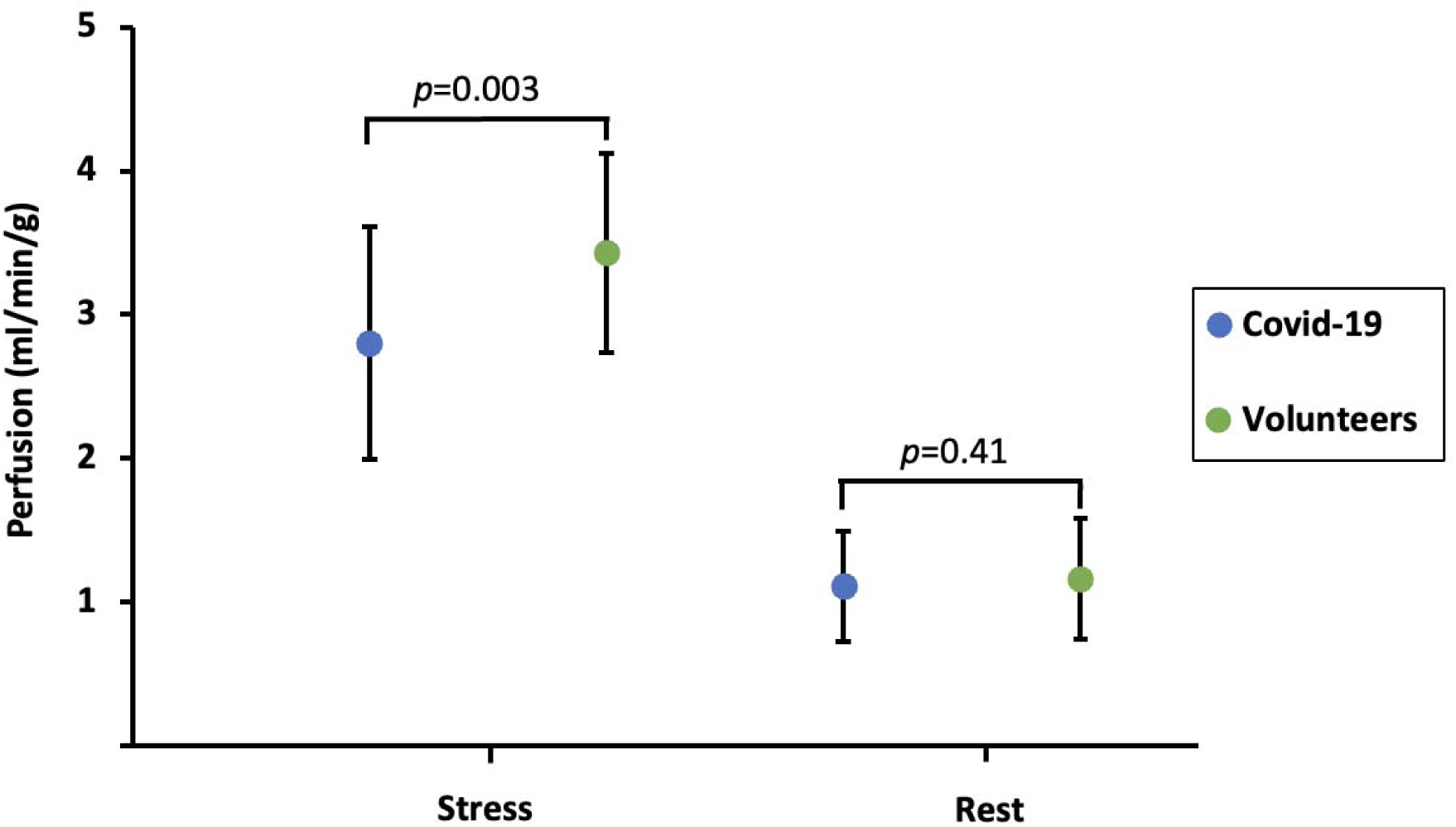
Stress and rest perfusion in COVID-19 patients and volunteers. The figure shows mean and standard deviation together with p-values. COVID-19 patients have lower stress perfusion but comparable rest perfusion, compared to volunteers.

**Figure 2.**
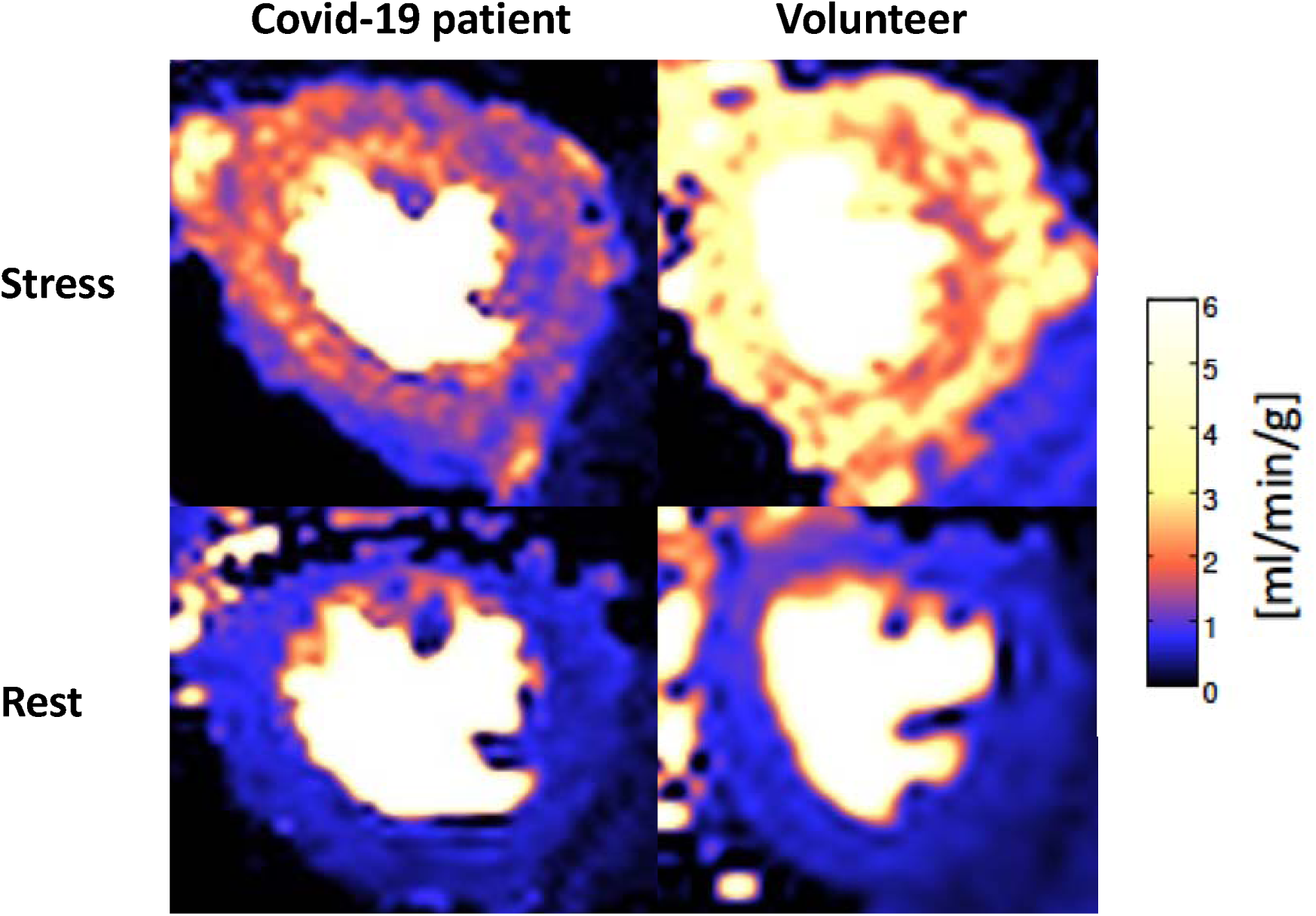
Midventricular perfusion maps of a COVID-19 patient (left) with suspected microvascular dysfunction and a volunteer (right), in stress (upper) and rest (lower). Rest perfusion is comparable, while stress perfusion is globally reduced in the COVID-19 patient compared to the volunteer.

## Discussion

To our knowledge, this is the first study evaluating possible CMD in long-term follow-up after hospitalization due to severe COVID-19, utilizing multiparametric CMR including quantitative adenosine stress perfusion mapping. Our study shows that COVID-19 patients from the first wave of the pandemic, with initially increased hs-TnT and/or PAP, 10 months later exhibit lower stress perfusion and a trend towards lower MPR, compared to volunteers without symptomatic IHD, Central Illustration. Additionally, COVID-19 patients had worse GLS and GCS, indicating impaired myocardial deformation, but volumes and LVEF did not differ. Minimal LGE was noted in four patients, but no focal perfusion defects. Notably, there were no differences in perfusion or MPR between COVID-19 patients with or without prior cardiovascular risk factors, or symptoms at follow-up.

### Conventional CMR in follow-up of COVID-19

Perfusion defects and predominantly non-ischemic LGE have been noted on CMR during acute COVID-19 or in early convalescence, with varying degrees of elevated native T1, native T2 and ECV, indicating inflammation, edema, and/or diffuse fibrosis^31^. Particularly, markers of systemic inflammation, native T1, native T2, and LGE have been shown to be higher in early convalescence following severe COVID-19 compared to mild or moderate cases, potentially due to inflammation-mediated cardiac injury^32^. Increased native T1 and native T2 in hospitalized COVID-19 patients may normalize within 6 months despite persistent cardiopulmonary symptoms^33,34^, while ECV may normalize within one year^35^. This is accordance with our study, where no differences were found for LV mass or volumes, native T1 or T2, nor ECV, between COVID-19 patients and volunteers 10 months post-discharge.

There is a discrepancy between long-term PACS symptoms and the relatively minor objective findings in previous cardiopulmonary evaluations^36^. In line with this, despite symptoms like chest pressure and dyspnea as possible indicators of heart failure, neither cardiac volumes nor LVEF differed between patients and volunteers. In COVID-19 patients with initial LV or RV functional and/or structural alterations, CMR a year later showed improved cardiac strain together with symptomatic improvement^37^. However, impaired GLS with normal LVEF has been shown 6 months after intensive care for COVID-19^38^. We showed impaired GLS and GCS at 10 months after intensive care for severe COVID-19, indicating some degree of persistently reduced cardiac function. Previous studies have shown that impaired GLS may be used, not only to evaluate cardiac improvement during follow-up of COVID-19, but also predict early mortality risk^39^.

### CMD and perfusion imaging in PACS

CMD may cause symptoms such as chest pain and dyspnea, but cannot be identified through conventional CMR, pulmonary radiology or functional testing. In early convalescence following COVID-19 with persistent cardiovascular symptoms, increased rest perfusion and/or reduced stress perfusion and MPR indicating CMD have been observed, even in mild Covid-19^40,41^. However, findings are inconsistent. Longitudinal studies using adenosine stress perfusion CMR in mild to moderate initial COVID-19 and persistent symptoms ^42,43^, and even with severe COVID-19 requiring hospitalization including intensive care^44,45^, have shown varying results regarding CMD. Alternative findings include focal inducible perfusion deficits and/or LGE with MI- or non-ischemic patterns ^44^ and mild myocarditis-like injury and signs of IHD, in many with no prior history of such^45^. Our study demonstrates reduced stress perfusion and a tendency towards lower MPR 10 months after severe COVID-19.

Quantitative CMR perfusion mapping has been validated against positron emission tomography (PET)^46^, which is reference standard for non-invasive quantitative perfusion imaging^47^. However, PET lacks the broader differential diagnostic capabilities of multiparametric CMR, which was employed in this study. Previous PET studies have indicated CMD in PACS by reduced stress perfusion and MPR at 5 months in hospitalized and non-hospitalized patients with ongoing symptoms^48^. At 11 months follow-up, combining PET with computed tomography (CT) to exclude CAD, MPR was decreased due to elevated rest perfusion, while stress perfusion was unaffected^49^. Importantly, reduced stress perfusion and MPR, identified via PET, were found in 50% of above middle-aged symptomatic males with cardiovascular risk factors at 6 months^50^. This finding was associated with increased risk of major adverse cardiovascular events including death at 10 months^50^. The MPR impairment was particularly pronounced following severe COVID-19, especially in patients admitted to intensive care^51^. Therefore, it is noteworthy that our study, which focuses on a predominantly male cohort of previously severely ill PACS patients, also found reduced stress perfusion and persistent cardiac symptoms at 10 months follow-up.

The variability in findings in previous studies of PACS patients in short- and long-term follow-up, can likely be attributed to differences in populations studied, initial disease severity, presence of cardiopulmonary symptoms at follow-up, and variations in timing and methodology of imaging^52^. It is surprising that, in contrast to the current study, most earlier studies have reported CMD predominantly in younger female patients with initial mild or moderate COVID-19. Our cohort mainly consisted of middle-aged males, many overweight but with a relatively low prevalence of known hypertension, type 2 diabetes, or hyperlipidemia. An association between CMD as determined by PET, and the metabolic syndrome, diabetes, and hypertension has been established^53^. Obesity is linked to CMD, as evidenced by lower stress perfusion and MPR, assessed by stress perfusion CMR^54^. Traditional cardiovascular risk factors associated with CMD also correlate with severe COVID-19^14^, complicating efforts to ascertain whether CMD results from severe COVID-19 or represents a pre-existing condition^55^. Although we did not study treatment effects in this study, our findings may inspire therapeutic aspects to be included in future studies of CMD.

## Limitations

This study is limited by its small sample size and single-center design. Our study represents the first long-term multiparametric CMR study, including quantitative stress perfusion mapping, in a unique cohort of first wave severely ill COVID-19 patients exhibiting long-term persistent cardiac symptoms. Moreover, we compared findings to age- and sex-matched volunteers. Little long-term data on CMD in this patient category exists and, although generalizability to the general PACS population may be limited, our results should be applicable for the proportion of new COVID-19 patients who become critically ill since they are unvaccinated or not optimally treated. Moreover, symptoms such as dyspnea, chest pressure, and fatigue may arise from respiratory and/or cardiovascular impairment^34^, but our study aim with a focus on cardiac parameters does not allow any interpretation of pulmonary contribution. Since patients were recruited prospectively after a clinical post-discharge follow-up evaluation, the amount of improvement in CMR parameters from the initial phase cannot be determined. Given prior studies indicating normalization at 6-12 months follow-up, it is conceivable that the reduced stress perfusion and impaired strain identified in this study represent residual effects of initially more severe cardiac involvement.

## Conclusions

COVID-19 patients exhibit long-term reduced stress perfusion indicating CMD, and impaired LV function by GLS and GCS. Lack of variation in stress perfusion between patients with and without cardiovascular risk factors suggests that CMD may be a consequence of severe COVID-19, warranting further investigation to elucidate mechanisms, and guide potential therapeutic interventions.

## Clinical Perspectives

### Competency in medical knowledge

Covid-19 patients with severe initial disease and persistent cardiac symptoms have signs of CMD and cardiac dysfunction in long-term follow-up using multiparametric stress perfusion CMR.

### Translational outlook

Future studies should explore, in larger groups in multi-center settings, the presence and pathophysiology of CMD and cardiac dysfunction in Covid-19 patients at index and over time, to establish a firm basis for therapeutic studies and secondary prevention.

## Data Availability

The data supporting the findings are available from corresponding author upon reasonable request.

## Abbreviations

ARDS: Acute Respiratory Distress Syndrome
CMD: Coronary Microvascular Dysfunction
CMR: Cardiovascular Magnetic Resonance Imaging
COVID-19: Coronavirus disease 2019
hs-TnT: High Sensitivity Troponin T
IHD: Ischemic Heart Disease
MPR: Myocardial Perfusion Reserve
PAP: Pulmonary Artery Pressure
PACS: Post-Acute COVID-19 Syndrome
PET: Positron Emission Tomography

---

**Central illustration.**
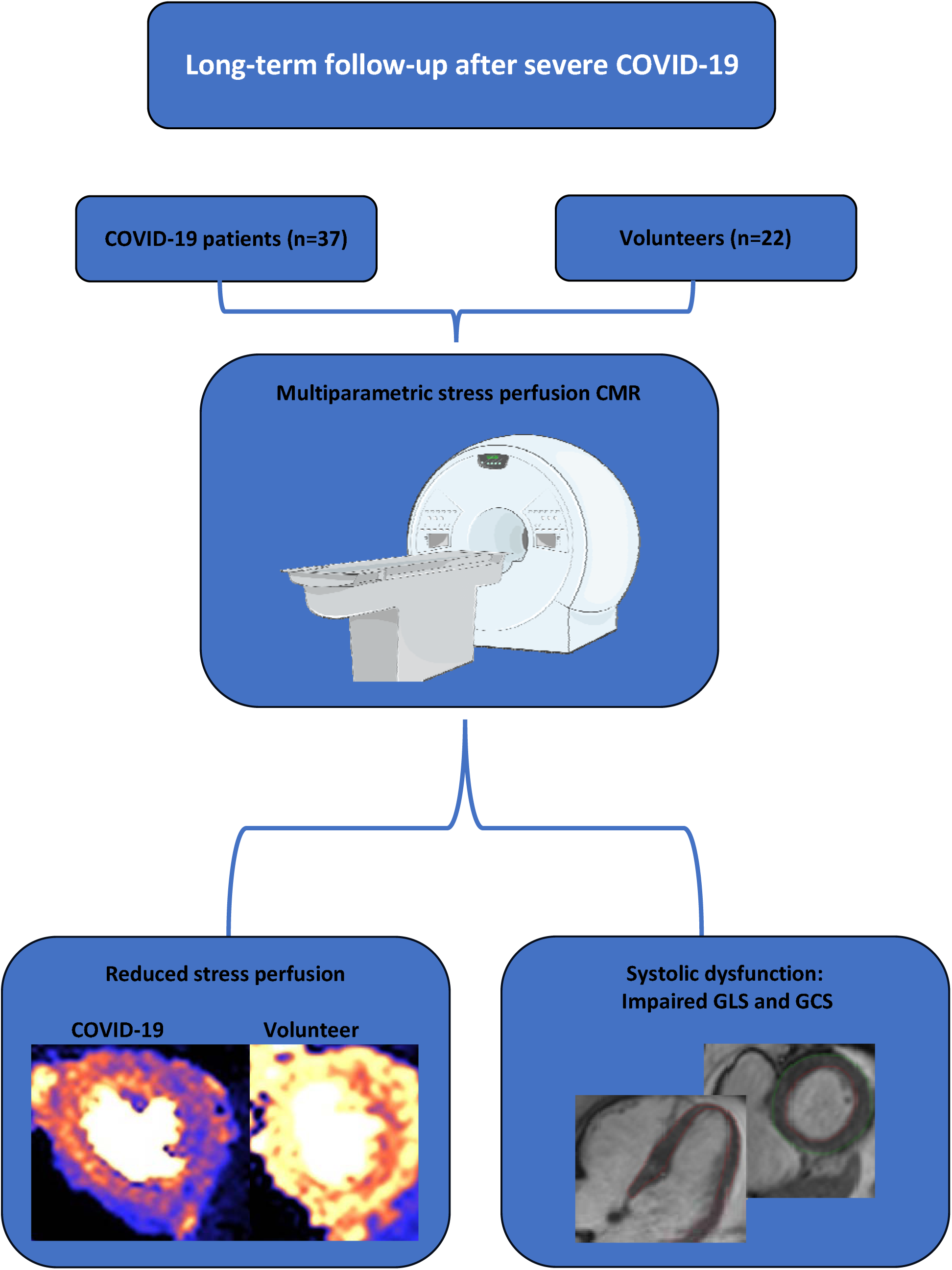
This follow-up study of hospitalized critically ill COVID-19 patients, using multiparametric stress perfusion CMR, showed reduced stress perfusion and impaired GCS and GLS at 10 months post-discharge, compared to volunteers of similar age and sex, without symptomatic IHD. Abbreviations: COVID-19 = Coronavirus disease 2019; GCS = global circumferential strain; GLS = global longitudinal strain; IHD = ischemic heart disease; CMR = cardiovascular magnetic resonance.

## Declarations

## Ethics, consent and permission

The study was approved by the Swedish Ethical Review Authority (Dnr 2021-03293, 2022-0695, 2020-02397). All patients provided written informed consent, including consent for publication of individual details on group level and anonymized images. Consent forms are held in the patient records and are available for review by the Editor-in-Chief. The data supporting the findings are available from corresponding author upon reasonable request.

## Authors contributions

RSJ participated in the design of the study, performed patient inclusion, analyzed images, and performed statistical analysis and drafted the manuscript. KL performed inter-observer analysis. DL participated in patient inclusion and image acquisition. KC, JB and MR participated in the study design, while JB and MR were also clinically responsible for all post-COVID-19 patients at Karolinska University Hospital. HX, PK participated in study design and developed the perfusion mapping technique. HE conceived the study, participated in study design and interpretation of data. JN participated in the design of the study, patient inclusion and image acquisition, and supervised all analysis, interpretation of data, and drafting of the manuscript. All authors read, revised and approved the final manuscript.

## Acknowledgements

We would like to acknowledge radiographer Elina Malkeshi and the biomedical scientist group for their invaluable work performing the image acquisition in this study. The Central Illustration includes a non-adapted figure provided by Servier Medical Art, licensed under a Creative Commons Attribution 4.0 Unported License (https://creativecommons.org/licenses/by/4.0/).

